# Semantic and geographical analysis of Covid-19 trials reveals a fragmented clinical research landscape likely to impair informativeness

**DOI:** 10.1101/2020.05.14.20101758

**Authors:** Giulia Tini, Bruno Achutti Duso, Federica Bellerba, Federica Corso, Sara Gandini, Saverio Minucci, Piergiuseppe Pelicci, Luca Mazzarella

**Affiliations:** Dept of Experimental Oncology, European Institute of Oncology IRCCS Milan; Department of Hemato-Oncology, University of Milan; Division of Early Drug Development for Innovative Therapies, IEO European Institute of Oncology IRCCS, Milan, Italy

## Abstract

**Background:** The unprecedented impact of the Covid-19 pandemics on modern society has ignited a “gold rush” for effective treatment and diagnostic strategies, with a significant diversion of economical, scientific and human resources towards dedicated clinical research. We aimed to describe trends in this rapidly changing landscape to inform adequate resource allocation.

**Methods:** We developed informatic tools (Covid Trial Monitor) to analyze in real time growth rate, geographical distribution and characteristics of Covid-19 related trials. We defined structured semantic ontologies with controlled vocabularies to categorize trial interventions, study endpoints and study designs. Data and analyses are publicly available at https://bioinfo.ieo.it/shiny/app/CovidCT

**Results:** We observe a clear prevalence of monocentric trials with highly heterogeneous endpoints and a significant disconnect between geographic distribution and disease prevalence, implying that most countries would need to recruit unrealistic percentages of their total prevalent cases to fulfill enrolment.

**Conclusions:** This geographically and methodologically incoherent growth sheds doubts on the actual feasibility of locally reaching target sample sizes and the probability of most of these trials providing reliable and transferable result. We call for the harmonization of clinical trial design criteria for Covid19 and the increased use of larger master protocols incorporating elements of adaptive designs. Covid Trial Monitor identifies critical issues in current Covid19-related clinical research and represents a useful resource for researchers and policymakers to improve the quality and efficiency of related trials

## Introduction

Standard and effective approaches for Covid19 prevention and treatment are not available to date, despite the magnitude of the pandemics and the similarities with past Coronavirus-associated diseases SARS and MERS[1]. So far initial trials with antivirals or other potentially effective drugs such as chloroquine have not yet clearly demonstrated superior efficacy over alternative treatments [2–4] and the disease remains associated with devastating morbidity and mortality. A wide variety of intervention strategies have been proposed, aiming for different mechanisms (viral or host processes), disease stage (early, advanced or prevention), intervention modality (medical or nonmedical).

As Covid-19-devoted resources grow, quantifying the potential impact of Covid19 trials becomes a relevant matter for global and national health policies. However, quality research on clinical trials is rendered difficult by the lack of standardized definition of trial parameters. Data reporting in trial repositories is notoriously plagued by internal inconsistencies, especially for “free text” fields that contain key information like inclusion criteria or study endpoints [5]. General medical ontologies like MeSH terms provide an all-encompassing framework but may be inadequate to capture relevant distinctions for specific fields; Covid-related terms were only introduced in late march, and their use is only recommended and not mandatory for trial definition.

In the present work we defined structured semantic ontologies with controlled vocabularies to categorize trial interventions, study endpoints and study designs, and we conducted an analysis of growth rate, geographical distribution and trial characteristics of Covid19-related trials, highlighting a number of relevant features which may impair the possibility of obtaining reliable and transferable results within the current framework. We formulate proposals for more rational trial designs against this rapidly changing landscape

## Results

### Global growth rate

We identified 1756 relevant studies (including interventional, observational and other) combining entries from the WHO and clinicaltrials.gov databases (figure 1A).

**Figure 1.**
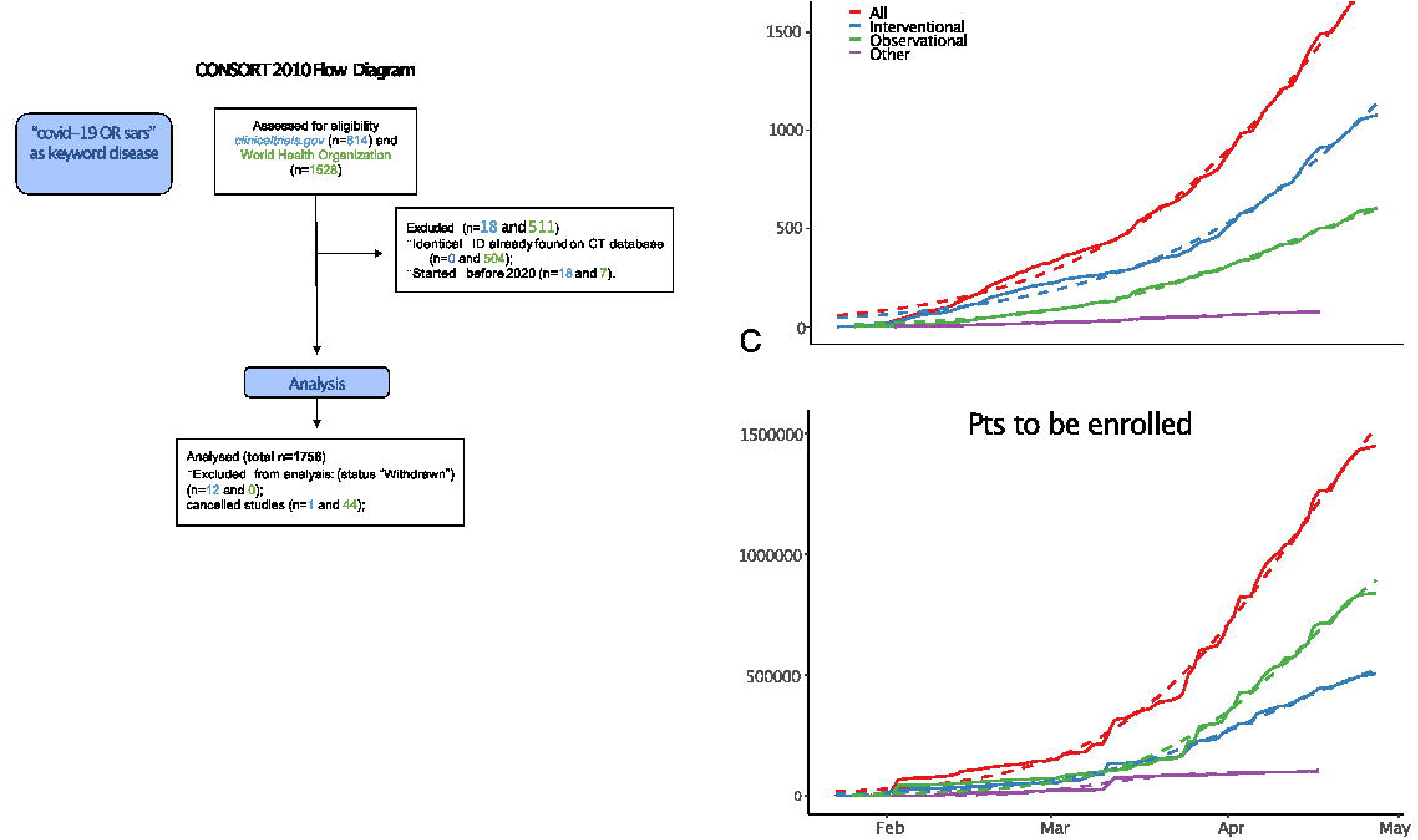
A. CONSORT diagram B. Cumulative growth of trials (top) and projected enrolled patients (thousands, bottom). See supplementary data for equation parameters

From 23 jan 2020 (date of first study posted), the cumulative increase of number of studies and the projected enrolled patients have been growing logistically (figure 1B).

We analysed the funding source on the 519 interventional trials from clinicaltrials.gov for which this information was available (table 1) and found a high percentage (397, 76.49%) are funded by public agencies, 62 (11.95%) by industries and 52 (10.02%) by collaboration among public and private. Comparison with a disease of comparable magnitude like influenza or cancer shows how this ratio of industry vs non-industry is highly unusual (influenza 15/47, 31.91%, p = 7.75×10^−5^, cancer 735/3167, 23.21%, p<2.2×10^−16^).

**Table 1.** Sources of funding.

| **STUDY**<br>**TYPE** | **INTERVENTIONAL** | **OBSERVATIONAL** | **TOTAL** |
| --- | --- | --- | --- |
| **OTHER** | 397 | 241 | 638 |
| **INDUSTRY** | 62 | 11 | 73 |
| **OTH/IND** | 52 | 7 | 59 |
| **OTH/NIH** | 3 | 0 | 3 |
| **NIH** | 2 | 5 | 7 |
| **U.S.FED** | 2 | 0 | 2 |
| **U.S.FED/OTH** | 1 | 0 | 1 |
| **TOTAL** | 519 | 264 | 783 |

Limited to clinicaltrials.gov. “OTH/IND” represent studies that were funded by a collaboration between industry and public sources, “OTH/NIH” studies funded by public sources and NIH, “U.S.FED/OTH” those funded by public sources and U.S. Fed.

For subsequent analyses, we focused on interventional trials (n = 1078), although data have been collected for all trials and are available in supplementary table 1 and at https://bioinfo.ieo.it/shiny/app/CovidCT.

### Geographical distribution

Trials were opened in 63 different countries. At the national level, United States was the nation with the highest number of trial locations, followed by China (figure 2A, C, D).

**Figure 2.**
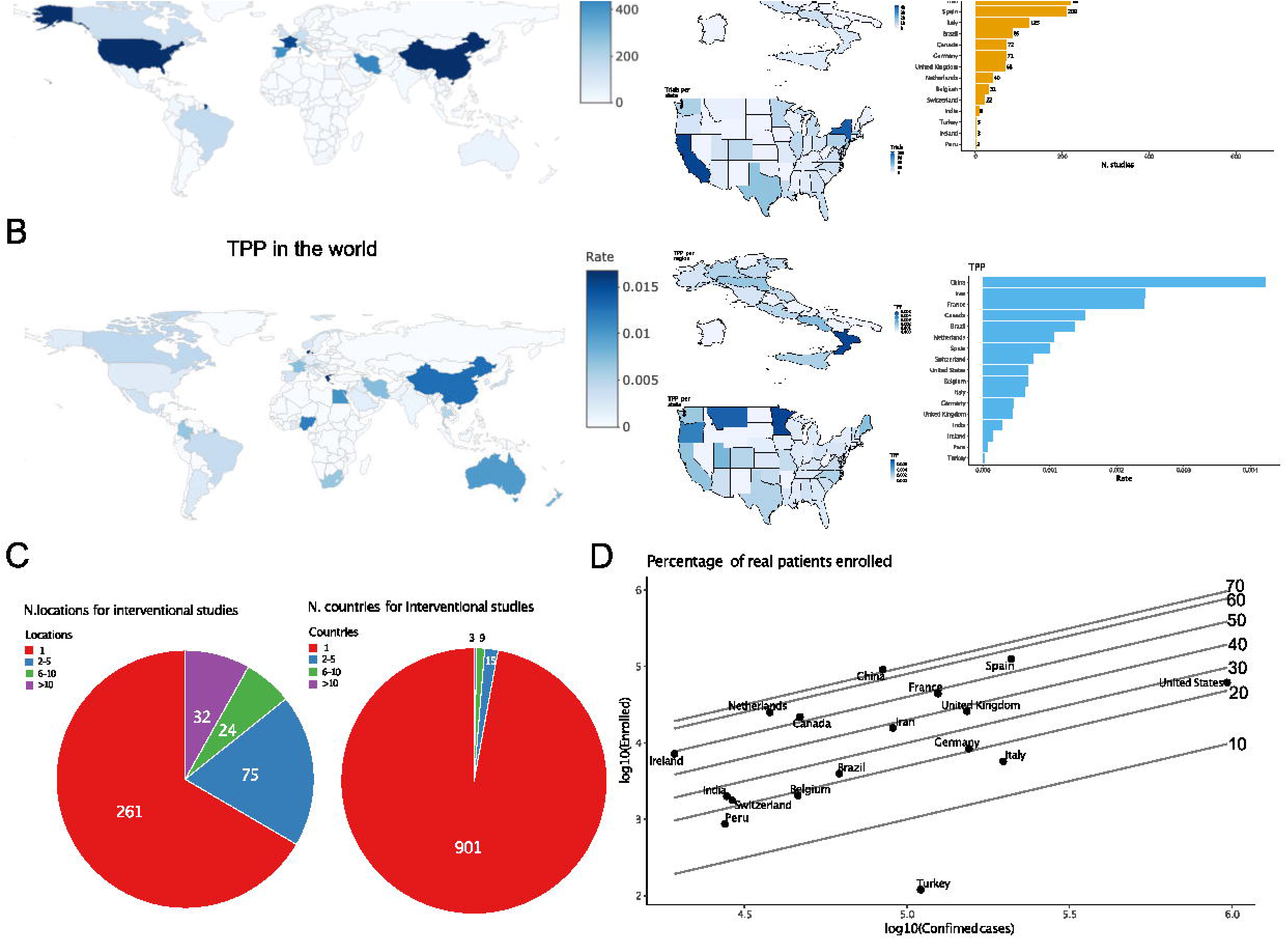
Geographical distribution. A. Total trials by nation (left), region or state in Italy and USA (middle), and bar graph of the first xxxx nations. Countries with <1000 confirmed cases are not reported B. Trial Per Patient (TPP) by nation (left), region or state in Italy and USA (middle), and bar graph of the first xxxx nations. Countries with <1000 confirmed cases are not reported C. Distribution of trials with 1, 2–5, 6–10 or > 10 locations (left) or states (right) D. Relationship between projected national enrolment and current cumulative confirmed cases, by state. Reference lines project the percentage of all confirmed cases to be enrolled. If a point sits on the 10% line, it means that 10% of all confirmed cases must be enrolled in a trial to satisfy enrolment projections for that country

We calculated a simple “trial per patient” index (TPP) by dividing the number of available trials by the number of cumulative cases. Trials per patient (TPP) were unevenly distributed among and within nations (figure 2B, E, F), with a Gini coefficient equal to 0.76. Of the 392 trials with available location information, the vast majority were monocentric (261, 66.58%), while 131 were multicentric. Of those, just 32 were opened in more than 10 locations (figure 2G).

The correlation between the cumulative projected patient enrolment and the actual case prevalence in each state was poor (Pearson = 0.37). With current case prevalence, most countries would need to recruit extremely high and possibly unrealistic percentages of their total prevalent cases to fulfill enrolment (figure 2H).

### Characteristics of interventional trials and types of intervention

Early phase studies (phase 1 and 1–2) were under-represented in both numbers and patients (figure 3A-B). To better describe and capture the semantic heterogeneity of trial characteristics, we defined ontologies with controlled vocabularies for interventions, study designs (supplementary table 2), inclusion criteria (supplementary table 3 and study endpoints (supplementary table 4).

**Figure 3.**
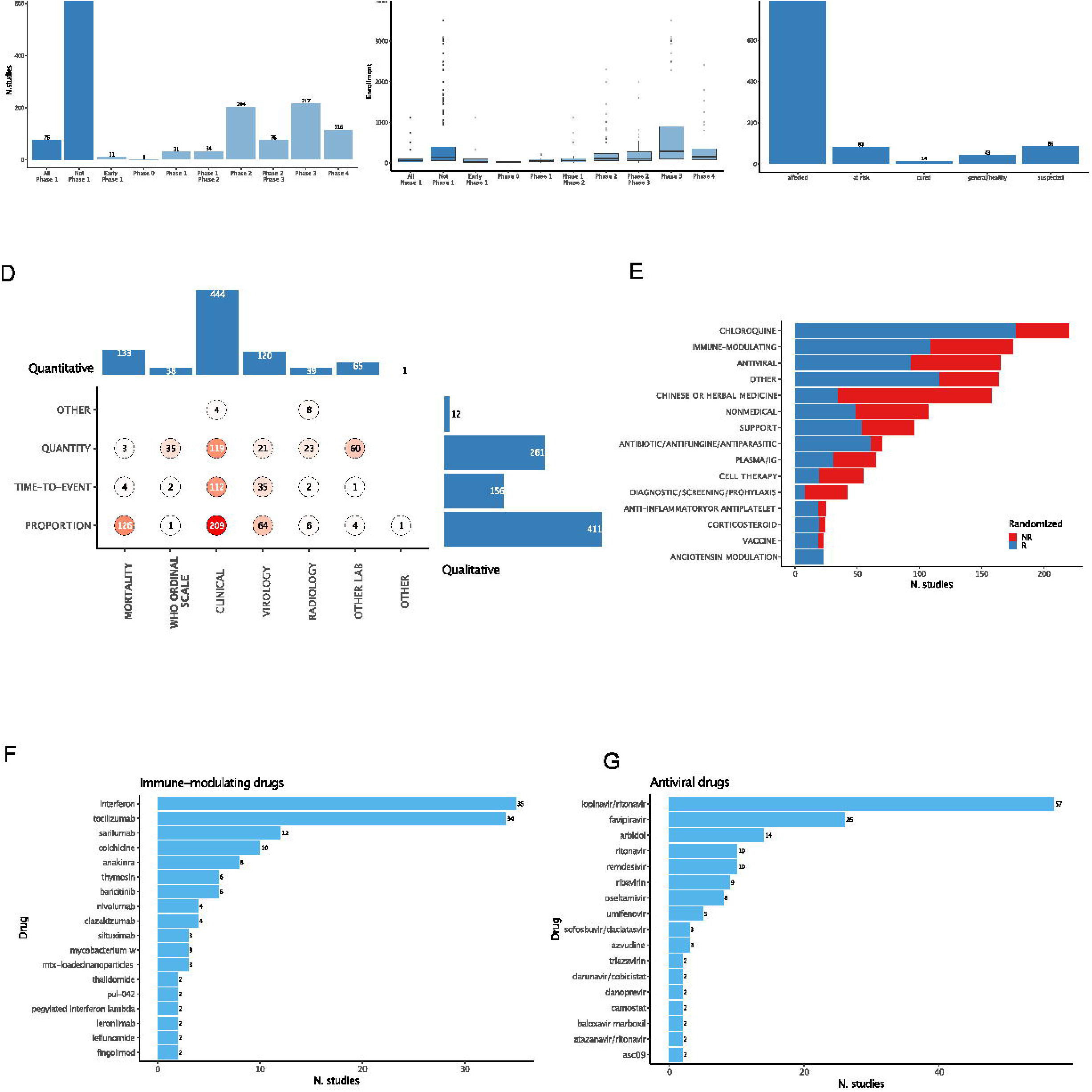
Trial features. A. Number of trials by phase. B. Cumulative planned enrolled patients by phase. One trial with > 4000 enrolment was removed from the graph C. Distribution of inclusion criteria D. Distribution of primary study endpoints. “Hard” endpoints, defined as including mortality, are in bold E. Distribution of intervention categories and use of randomized designs F. Breakdown of immune-modulating drugs G. Breakdown of antiviral drugs

Among trials aimed at active treatment, a significant share (86/1078) do not require PCR-confirmed diagnosis as inclusion criteria (“suspected”, figure 3C). Primary endpoints are qualitatively (clinical or virological, radiological or other laboratory variables) and quantitatively (411 proportion, 156 time-to-event, 261 quantity) heterogeneous; “hard” endpoints containing mortality either use the incommensurable quantitative WHO ordinal scale or proportional measures (figure 3D).

We categorized all interventional treatments under 15 terms. Randomization is common but not prevalent among most interventions (figure 3E); Chloroquine, immune-modulating agents (expanded in figure 3F) and antivirals (expanded in fig 3G) are the most investigated with 220, 175, and 165 studies respectively.

## Discussion

We present quantitative, updated and semantically organized measures of Covid19-related trials. We highlight a number of peculiar characteristics of this clinical research landscape: extremely rapid growth, substantial geographical and methodological incoherence, unprecedented funding pattern, prevalence of monocentric trials, extreme heterogeneity in the interventions tested. The main limitation of our analysis is represented by the heterogeneity in quality and quantity of available information. The source databases use often non-overlapping trial categorization methods and many of the records have missing, mis-spelled or imprecise wording, potentially causing relevant selection biases which we attempted to mitigate by forcing information through controlled vocabularies, a procedure which may results in loss of information.

We argue that several of the planned trials are unlikely to provide high-quality results for the following reasons.

First and foremost, the unrealistic percentages of total prevalent cases needed to fulfill planned enrollment at the national level implies that several trials are unlikely to reach target sample sizes, with severe loss of statistical power or study termination. This has in fact already been observed with the recently published Remdesivir trial in China, which failed to complete enrolment and led to conflicting interpretations[6].

Geographical fragmentation will magnify local and study-specific confounding in demographics, comorbidities and the availability of healthcare resources, which are known to heavily impact on Covid-19 outcome [7–9]. Variegated endpoints and inclusion criteria will inhibit the possibility to adequately compare and meta-analyse treatments across trials. Proper dose-finding trials are scarce, with the risk of under- or over-treating patients and of underestimating potentially risky drug-interactions. Finally, the scientific soundness of classical randomized designs in a scenario where the control arm may be rapidly changing[10] is ethically and methodologically questionable Our analysis provides quantitative grounds for concerns raised in the early days of the Covid19 pandemics in commentaries [11–13] that highlighted the difficulties in striking a balance between the need to conduct sound clinical research and the need to rapidly take action. This observed disordered growth in clinical research is perhaps expected given the unprecedented medical and socio-economical impact of the Covid-19 pandemics. However, we note that the scientific community should prepare the grounds for a more ordered development, especially in light of the expected persistence of SARS-CoV2 and the likely emergence of other Coronavirus-mediated diseases in the long run.

A potential solution for some of the above issues is to favour the adoption of adaptive trial design features (inclusion of predefined toxicity/efficacy stopping rules, biomarker-adjusted enrolment, etc) and the inclusion of multiple phases, interventions and patient groups under the same regulatory framework, using the so-called “master protocol” model[14]. Advantages of this model include: i) the possibility to compare efficacy of multiple interventions against a single, well-standardized control arm ii) the possibility to compare across multiple treatments, particularly relevant in a scenario where time bias is likely to play a major role: mortality is likely to be subject to time-dependent variables such as the ICU occupancy ratio or acquired physician experience iii) the possibility to skew enrolment into more effective/less toxic arms as new data are accumulated iv) the possibility to introduce novel treatment arms or stratification biomarkers as these are identified in preclinical or translational studies v) the possibility to collect samples for translational studies under a unified and homogeneous framework, increasing their informativeness. We identified 18 trials with declared adaptive features in their designs (supplementary table 5), among which most notable is the large SOLIDARITY trial promoted by WHO to test 4 treatment options (Remdesivir; Lopinavir/Ritonavir; Lopinavir/Ritonavir with Interferon beta-1a; and Chloroquine or Hydroxychloroquine) against standard of care.

Master protocols are themselves subject to specific biases, in particular the need to adjust for multiple hypotheses testing [14, 15], and often require sophisticated monitoring and logistics which can only be accomplished within large organizations. This calls for stronger interaction between stakeholders like pharma companies, regulatory bodies, funding entities and patient organizations. In the present rapidly changing scenario such frameworks may be of particular utility to efficiently discard nonviable hypotheses and prioritize treatment that deserve proper testing on larger scales. Experience gained in some fields where master protocols are increasingly adopted, such as oncology [16, 17], may inform trial design.

## Methods

### Databases

Data were downloaded from clinicaltrials.gov and the WHO International Clinical Trials Registry

Platform (ICTRP https://www.who.int/ictrp/en/) on April 11^th^ and 27^th^

Data for Covid cases by country and for US states were downloaded from Johns Hopkins Data

Repository (https://github.com/CSSEGISandData/COVID-19) and for Italian regions from Presidenza del Consiglio dei Ministri – Dipartimento della Protezione

Civile (https://github.com/pcm-dpc/COVID-19) on April 27th

Details on ontology definition, geographical analyses and statistical analyses are discussed in supplementary methods

## Data Availability

data are made available at https://bioinfo.ieo.it/shiny/app/CovidCT

## Acknowledgments

We thank Dr Arnaud Ceol for informatic support

